# Alignments between cortical neurochemical systems, proteinopathy and neurophysiological alterations along the Alzheimer’s disease continuum

**DOI:** 10.1101/2024.04.13.24305551

**Authors:** Alex I. Wiesman, Jonathan Gallego-Rudolf, Sylvia Villeneuve, Sylvain Baillet, Tony W. Wilson, the PREVENT-AD Research Group

**Author notes:** Corresponding author, Correspondence: Alex I. Wiesman, PhD, McConnell Brain Imaging Centre Montreal Neurological Institute McGill University, Montreal QC. Authors contributed equally to this report (Sylvain Baillet, Tony W. Wilson). Data used in preparation of this article were obtained from the Pre-symptomatic Evaluation of Novel or Experimental Treatments for Alzheimer’s Disease (PREVENT-AD) program (https://douglas.research.mcgill.ca/stop-ad-centre). A complete listing of PREVENT-AD Research Group can be found in the PREVENT-AD database: https://preventad.loris.ca/acknowledgements/acknowledgements.php?date=[2024-04-02]. The investigators of the PREVENT-AD program contributed to the design and implementation of PREVENT-AD and/or provided data but did not participate in analysis or writing of this report.

## Abstract

Two neuropathological hallmarks of Alzheimer’s disease (AD) are the accumulation of amyloid-β (Aβ) proteins and alterations in cortical neurophysiological signaling. Despite parallel research indicating disruption of multiple neurotransmitter systems in AD, it has been unclear whether these two phenomena are related to the neurochemical organization of the cortex. We leveraged task-free magnetoencephalography and positron emission tomography, with a cortical atlas of 19 neurotransmitters to study the alignment and interactions between alterations of neurophysiological signaling, Aβ deposition, and the neurochemical gradients of the human cortex. In patients with amnestic mild cognitive impairment (N = 18) and probable AD (N = 20), we found that changes in rhythmic, but not arrhythmic, cortical neurophysiological signaling relative to healthy controls (N = 20) are topographically aligned with cholinergic, serotonergic, and dopaminergic neurochemical systems. These neuro-physio-chemical alignments are related to the severity of cognitive and behavioral impairments. We also found that cortical Aβ plaques are preferentially deposited along neurochemical boundaries, and mediate how beta-band rhythmic cortical activity maps align with muscarinic acetylcholine receptors. Finally, we show in an independent dataset that many of these alignments manifest in the asymptomatic stages of cortical Aβ accumulation (N = 33; N = 71 healthy controls), particularly the Aβ-neurochemical alignments (57.1%) and neuro-physio-chemical alignments in the alpha frequency band (62.5%). Overall, the present study demonstrates that the expression of pathology in pre-clinical and clinical AD aligns topographically with the cortical distribution of chemical neuromodulator systems, scaling with clinical severity and with implications for potential pharmacotherapeutic pathways.

## Introduction

Alzheimer’s disease (AD) is marked by cortical accumulations of cytotoxic proteins including amyloid-β (Aβ) and hyper-phosphorylated tau, which are associated with alterations of neurophysiological activity^1–4^. These alterations are expressed across multiple frequency bands^1,2,5–11^, and are associated with the severity of hallmark impairments in memory, attention and executive function^1,5,12,13^. Unlike superficially similar relationships observed in Parkinson’s disease^14^, pathological alterations in AD are predominantly *rhythmic* in nature^15^. Rhythmic neurophysiology can be modulated non-invasively via frequency-targeted neuromodulation^16–21^, making aberrant neurophysiological oscillations a promising target for emerging therapeutics. Despite these advances, the mechanistic bases of neurophysiological alterations in AD are understudied in human participants.

Decades of research have also indicated that a subset of neurotransmitter systems are degraded as patients progress along the AD continuum. In particular, cholinergic and glutamatergic signaling are dysfunctional in AD with relevance for cognitive impairments^22–25^. Both of these neurotransmitter systems also represent pharmacological targets in AD, via acetylcholinesterase inhibitors and memantine, respectively^22,25^. More recently, losses of function in dopaminergic and serotonergic pathways have also been linked to psychiatric and behavioral symptoms in AD^26–29^. Due to their shared associations with cognitive and behavioral impairments, it is possible that the aforementioned neurophysiological alterations and accumulations of proteinopathy seen in patients with AD are influenced by these neurochemical systems.

Recent research has indicated that the topographical alignment of altered neurophysiological activity and neurochemical systems are clinically-informative in Parkinson’s disease^30^. However, similar relationships between neurochemistry and clinically-relevant alterations in macro-scale neurophysiology have not been characterized in patients with AD. In the current study, we examine the alignment of AD-related neurophysiological alterations and proteinopathy with the principal neurochemical systems of the cortex, as well as the associations between these alignments and the hallmark cognitive and behavioral symptoms of the disease. From the above review of the field, we hypothesized that rhythmic neurophysiological alterations in AD would align preferentially with cortical cholinergic, glutamatergic, dopaminergic and serotonergic systems, due to their shared involvement in the disease process. We expected that the extent of these alignments would relate to the severity of cognitive and behavioral symptoms. We also predicted that these alignments would be mediated by the regional deposition of Aβ plaques, indicating a stronger cytotoxic effect on neurophysiology in brain regions with specific neurochemical profiles. Finally, we hypothesized that these alignments would be detectable in asymptomatic older adults with high levels of cortical Aβ deposition, possibly indicating promising novel targets for early-stage disease monitoring and intervention.

## Results

### Topographies of neurochemical gradients and neurophysiological alterations & amyloid-β deposition in patients with aMCI and AD

**Figure 1.**
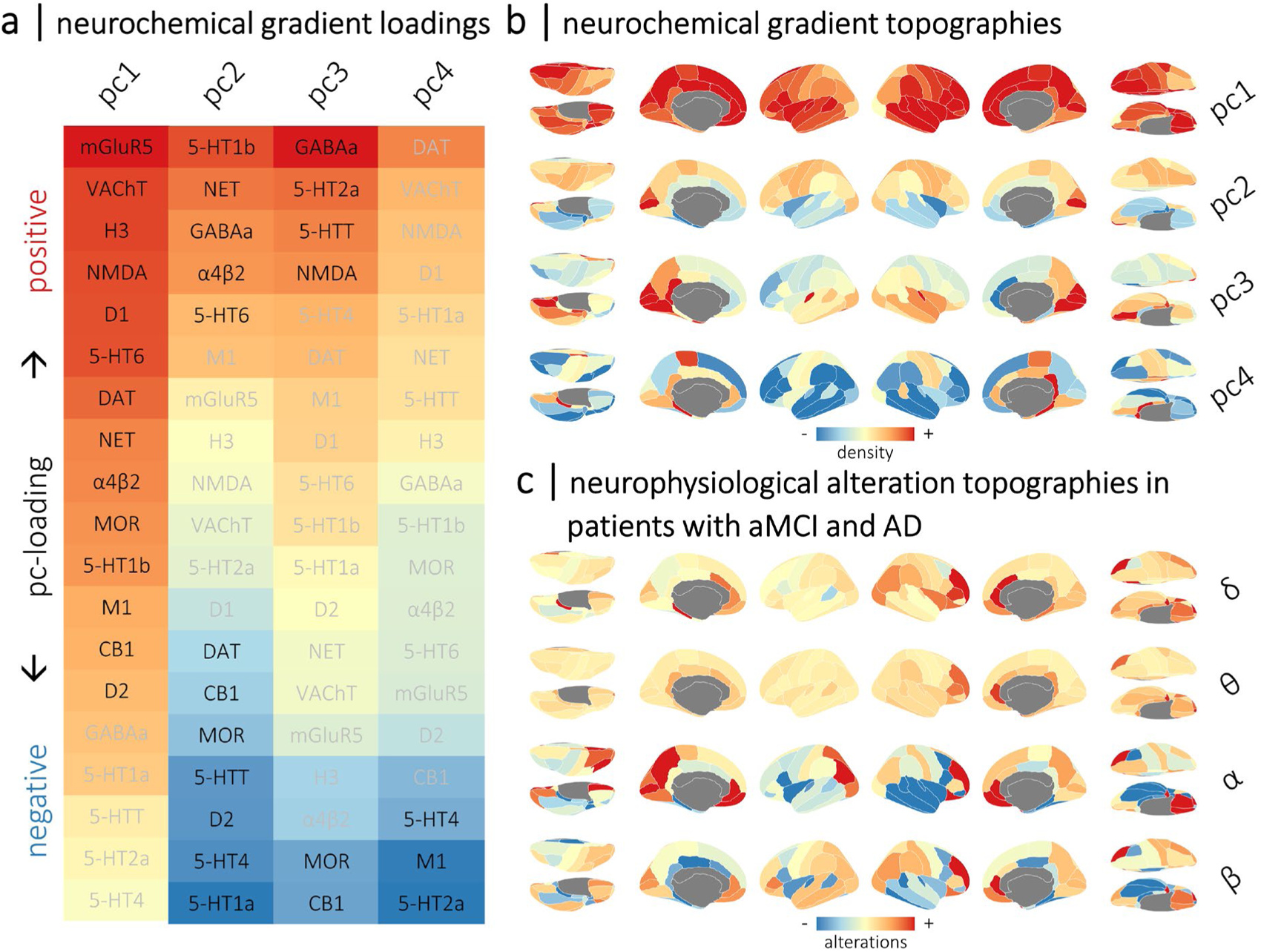
Topographies of normative neurochemical gradients and neurophysiological alterations in patients with aMCI and AD. Normative gradients of cortical neurotransmitter system density were derived from 19 normative neurotransmitter atlases using principal component analysis. The first four principal components (pc1 to pc4) explained more than 80% of the variance across atlases (all *p*’s < .001; 1,000 permutations). Heatmaps in (a) show the respective loadings (i.e., eigenvectors) for each neurochemical gradient across the 19 atlases. Positive and negative loadings are colored red and blue, respectively, and labels in black indicate systems that significantly contribute to each gradient (*p*’s < .05; 1,000 permutations). The spatial topographies of these neurochemical gradients are shown as cortical maps in (b), with warm colors indicating regions with high densities of positively-loaded neurochemical systems and cool colors indicating regions with high densities of negatively-loaded systems. The cortical maps shown in (c) indicate the group-average spatial topographies of alterations in rhythmic neurophysiology in patients with aMCI and AD from the DMAP cohort, per each canonical frequency band (i.e., δ: 2 – 4 Hz; θ: 5 – 7 Hz; α: 8 – 12 Hz; β: 15 – 29 Hz). Note that these topographies represent rhythmic neurophysiological measures that have been standardized (i.e., z-scored) to the comparable data from the healthy control participants. Warm and cool colors indicate increases and decreases in rhythmic activity relative to healthy levels, respectively.

To account for the complex co-localization and co-release of neurotransmitter systems in the human cortex, we data-reduced the 19 neurotransmitter system atlases into four neurochemical gradients using principal component analysis (1,000 permutations; accounting for 81.7% of total spatial variance; all *p*’s < .001). These gradients have been discussed extensively in previous work^30,66^ and are shown in Figures 1a and 1b. Briefly, the first gradient (pc1; eigenvalue = 6.13; 32.3% of spatial variance) exhibited positive loadings from most modeled receptors and transporters; the second gradient (pc2; eigenvalue = 4.79; 25.2% of spatial variance) contrasted cortical regions rich in norepinephrine, GABA, acetylcholine, and 5-HT1b/5-HT6 serotonin systems from those with higher densities of dopamine, mu-opioid, cannabinoid, and other serotonergic (5-HT1a, 5-HT4, and 5-HTT) systems; the third gradient (pc3; eigenvalue = 2.56; 13.5% of spatial variance) described a spatial pattern of regions rich in GABAergic, serotonergic (5-HT2a and 5-HTT), and glutamatergic (NMDA) systems versus those with lesser mu-opioid and cannabinoid systems; and the fourth gradient (pc4; eigenvalue = 2.04; 10.7% of spatial variance) mapped onto regions with low serotonergic (5-HT2a and 5-HT4) and acetylcholinergic (M1) receptor densities.

Cortical maps representing the mean region-wise alterations in rhythmic and arrhythmic neurophysiology across the DMAP patients with aMCI and AD are shown in Figures 1c and S1, respectively. These patients exhibited increases in low-frequency (i.e., delta- and theta-band) rhythms relative to healthy levels in prefrontal and parieto-occipital regions, alongside pronounced decreases in high-frequency (i.e., alpha- and beta-band) neurophysiological rhythms strongest in frontal and temporal areas. Beta rhythms were also selectively increased in parietal and prefrontal regions.

Patients with aMCI and AD exhibited increased aperiodic offsets and steeper aperiodic slopes in prefrontal and parieto-occipital regions relative to healthy participants, indicating increased arrhythmic signaling, particularly in slower frequencies (Figure S1).

The deposition of Aβ plaques also exhibited a stereotyped topography in patients with aMCI and AD: SUVRs were highest in prefrontal and parieto-temporal cortices bilaterally (Figure S1).

### Neurophysiological alterations in patients with aMCI and AD align with neurochemical systems

We tested the alignment of neurophysiological alterations in patients with aMCI and AD with the four neurochemical gradients using nested linear models (Figure 2; see *Methods: Modeling of Spatial Colocalization*).

Generally, AD-related alterations in rhythmic, but not arrhythmic, neurophysiological activity were aligned with neurochemical systems. Disease-related increases in slower delta and theta rhythms were stronger in cortical regions with low density of dopaminergic, serotonergic, and mu-opioid receptors/transporters, while disease-related decreases in faster alpha and beta activity were weaker in these regions. Alpha and beta rhythmic decreases were also stronger in brain regions with high densities of glutamatergic receptors, and increases in beta rhythms favored areas rich in muscarinic cholinergic systems.

Rhythmic activity in the slower delta (*t* = 4.34, *p*_FDR_ < .001) and theta (*t* = 3.16, *p*_FDR_ = .005) bands was aligned with the second principal neurochemical gradient (Figure 2a-b). For delta alterations, the neurotransmitter systems that contributed most to this colocalization were dopaminergic (DAT: *t* = -4.25, *p*_FDR_ < .001; D2: *t* = -3.80, *p*_FDR_ < .001), serotonergic (5-HT1a: *t* = -4.53, *p*_FDR_ < .001; 5-HT4: *t* = -4.23, *p*_FDR_ < .001), and mu-opioid (MOR: *t* = -4.36, *p*_FDR_ < .001). For theta rhythmic alterations, noradrenergic (NET: *t* = -3.81, *p*_FDR_ = .002), serotonergic (5-HT1a: *t* = -3.41, *p*_FDR_ = .004; 5-HT6: *t* = -2.96, *p*_FDR_ = .012), dopaminergic (D2: *t* = -2.75, *p*_FDR_ = .018), and GABAergic (GABAa: *t* = -2.47, *p*_FDR_ = .032) systems were aligned.

**Figure 2.**
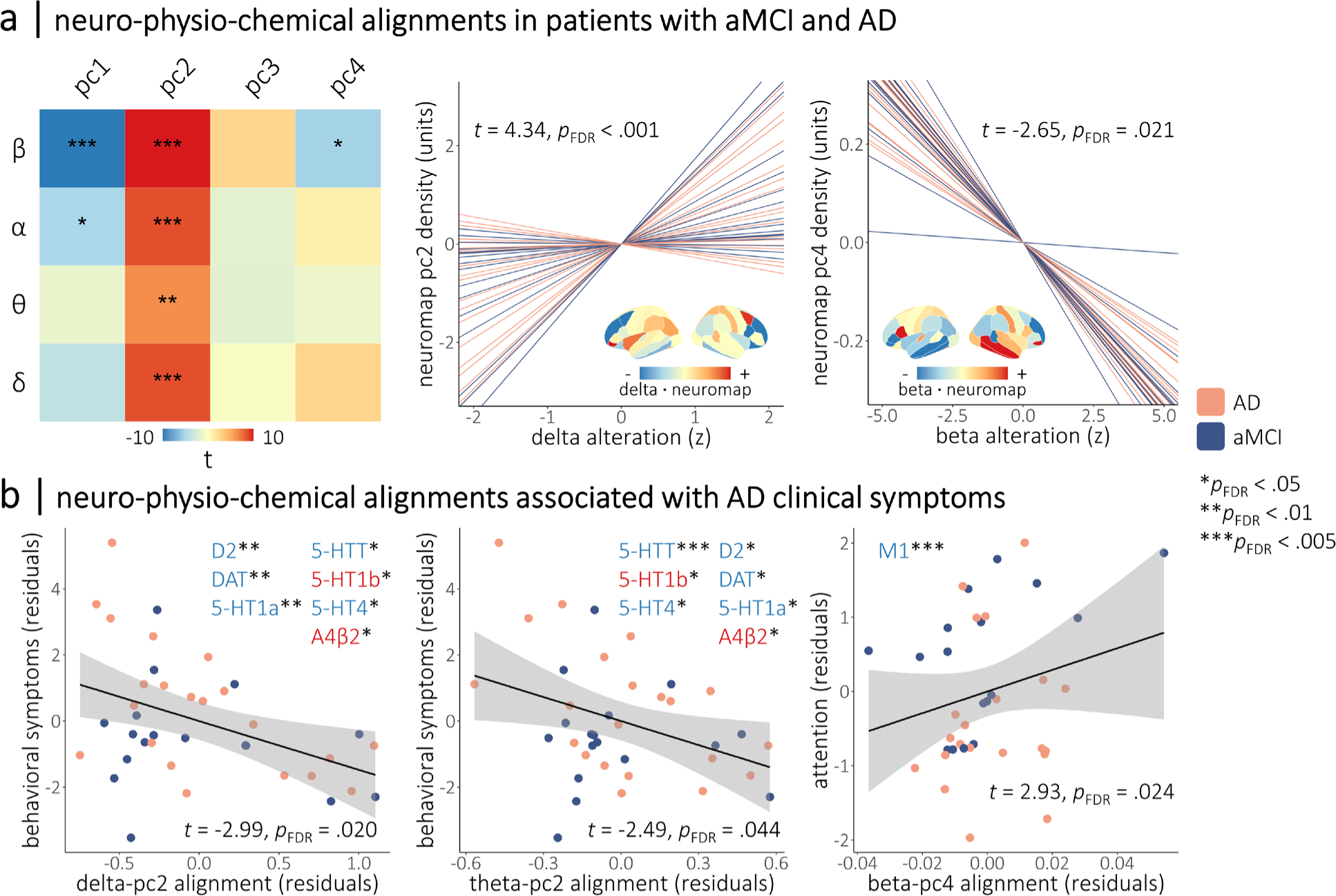
Neurophysiological alterations in patients with aMCI and AD align to neurochemical systems with relevance for clinical symptoms. The heatmap in (a) indicates the strength of the alignments between band-limited alterations in rhythmic neurophysiology (y-axis) and the normative neurochemical gradients (x-axis) from Figure 1. Colors indicate the t-values of these relationships, with asterisks indicating statistical significance after correcting for multiple comparisons. Line plots in (a) show the participant-level simple slopes representing two of these alignments, with the remaining alignments displayed in Figure S2. Overlaid cortical maps indicate the dot product of the relevant neurophysiological alterations and neurochemical gradient densities per region, with warm and cool colors indicating positive and negative agreement, respectively, between the two measures. Representative topographies of delta rhythmic alterations from patients with high and low alignment to the second neurochemical gradient are shown in Figure S3. Scatterplots in (b) indicate the moderations of neurochemical-neurophysiological alignments from (a) by cross-sectional clinical symptom severity, with the strength of alignment shown on the x-axis and the clinical symptom score shown on the y-axis. Inlaid neurotransmitter receptor/transporter abbreviations indicate the respective atlases that replicate the relevant moderation effect in post-hoc testing, with the color and saturation of each label indicating the sign of the corresponding loading onto the neurochemical gradient. For all plots, the color of the lines/points indicates whether patients were in the amnestic mild cognitive impairment or probable Alzheimer’s disease clinical subgroup. \*\*\**p*_FDR_ < .005, \*\**p*_FDR_ < .01, \**p*_FDR_ < .05.

We found that AD-related alterations in rhythmic alpha (*t* = -2.53, *p*_FDR_ = .026) and beta (*t* = -4.99, *p*_FDR_ < .001) activity colocalized negatively with the first neurochemical gradient (Figure 2a-b), with strongest alignments to dopaminergic (DAT-beta: *t* = -6.28, *p*_FDR_ < .001; DAT-alpha: *t* = -4.82, *p*_FDR_ < .001; D1-beta: *t* = -5.70, *p*_FDR_ < .001; D1-alpha: *t* = -2.93, *p*_FDR_ = .011; D2-beta: *t* = -5.44, *p*_FDR_ < .001; D2-alpha: *t* = -4.89, *p*_FDR_ < .001), mu-opioid (MOR-beta: *t* = -5.72, *p*_FDR_ < .001; MOR-alpha: *t* = -2.89, *p*_FDR_ = .011), and glutamatergic (NMDA-beta: *t* = -4.99, *p*_FDR_ < .001; NMDA-alpha: *t* = -3.32, *p*_FDR_ = .004) systems. These alterations in fast alpha and beta rhythms were also aligned positively with the second neurochemical gradient, again with strong contributions from dopaminergic and mu-opioid systems, and additional contributions from serotonin systems (5-HT1a-beta: *t* = -7.17, *p*_FDR_ < .001; 5-HT1a-alpha: *t* = -5.43, *p*_FDR_ < .001; 5-HTT-beta: *t* = -5.60, *p*_FDR_ < .001; 5-HTT-alpha: *t* = -3.16, *p*_FDR_ = .004; 5-HT4-alpha: *t* = -3.79, *p*_FDR_ < .001). Finally, alterations in beta rhythms were aligned negatively with the fourth neurochemical gradient (*t* = -2.65, *p*_FDR_ = .021), with contributions from serotoninergic (5-HT4: *t* = -4.98, *p*_FDR_ < .001) and cholinergic (M1: *t* = 3.94, *p*_FDR_ < .001) systems.

None of these alignments were moderated by clinical subgroup (i.e., aMCI versus AD). The alignments to the first neurochemical gradient remained significant when an atlas of synaptic density (i.e., glycoprotein) was included as a nuisance covariate, indicating they are not attributable to regional variations in non-specific receptor densities. No significant alignments to the third neurochemical gradient were observed. Arrhythmic neurophysiological alterations were not aligned to any of the four neurochemical gradients (all *p*_FDR_’s > .150). These arrhythmic models remained non-significant when the region-wise aperiodic model fits (standardized to those of the healthy control data) were included as a nuisance covariate.

### Neurophysiological-neurochemical alignments are associated with cognitive and behavioral symptoms

We examined whether the strength of these neurochemical alignments was associated with cognitive and behavioral symptoms in patients with aMCI and AD (Figure 2c).

We found moderations of the slow-rhythmic alignments to the second neurochemical gradient by behavioral symptoms (i.e., QDRS behavioral score; delta-pc2: *t* = -2.99, *p*_FDR_ = .020; theta-pc2: *t* = -2.49, *p*_FDR_ = .044), such that greater delta and theta rhythmic increases in brain regions rich in dopaminergic and serotonergic systems were related to worse behavioral symptoms.

We also found that the alignment between beta rhythmic alterations and the fourth neurochemical gradient was moderated by attention function (*t* = -2.93, *p*_FDR_ = .024), such that increased beta rhythmic activity, relative to healthy levels, in brain regions rich in muscarinic acetylcholine receptor M1 was related to worse cognitive abilities.

### Deposition of amyloid-β in patients with aMCI and AD aligns with neurochemical systems

We next tested whether the deposition of Aβ plaques in the DMAP participants occurs preferentially in brain regions with specific neurochemical profiles. We found alignment between the topographies of Aβ deposition and the second (*t* = 11.84, *p*_FDR_ < .001), third (*t* = -24.21, *p*_FDR_ < .001), and fourth (*t* = -16.22, *p*_FDR_ < .001) neurochemical gradients (Figure 3). The alignment of Aβ with the second neurochemical gradient was moderated by clinical subgroup, such that patients with a diagnosis of probable AD exhibited stronger alignments than those with aMCI (*t* = 2.49, *p*_FDR_ = .038).

These relationships were such that Aβ tended to accumulate in brain regions rich in cholinergic (α4β2: *t* = 20.22, *p*_FDR_ < .001; M1: *t* = 8.52, *p*_FDR_ < .001), cannabinoid (CB1: *t* = 9.23, *p*_FDR_ < .001), mu-opioid (MOR: *t* = 8.40, *p*_FDR_ < .001), noradrenergic (NET: *t* = 2.71, *p*_FDR_ = .007), and two of the serotonergic (5-HT6: *t* = 4.39, *p*_FDR_ < .001; 5-HT1b: *t* = 9.92, *p*_FDR_ < .001) systems, and conversely spared brain regions rich in dopaminergic (D2: *t* = -7.10, *p*_FDR_ < .001; DAT: *t* = -19.24, *p*_FDR_ < .001), glutamatergic (NMDA: *t* = -12.55, *p*_FDR_ < .001), GABAergic (GABAa: *t* = -8.18, *p*_FDR_ < .001), and other serotonergic (5-HT1a: *t* = -14.72, *p*_FDR_ < .001; 5-HT4: *t* = -9.64, *p*_FDR_ < .001; 5-HTT: *t* = -22.14, *p*_FDR_ < .001) systems.

**Figure 3.**
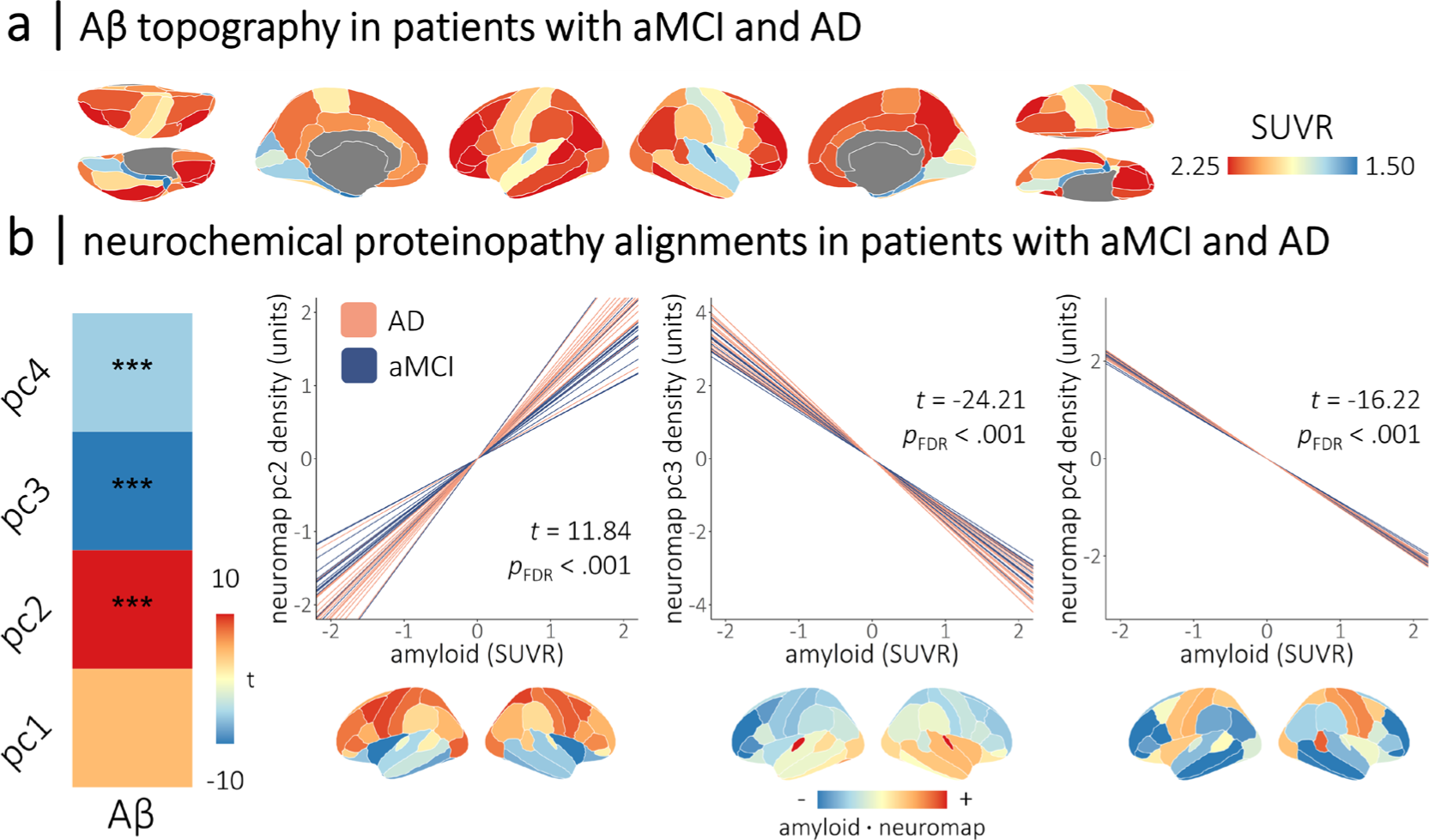
Cortical deposition of amyloid-β in patients with aMCI and AD aligns with neurochemical systems. Cortical maps in (a) show the mean topography of amyloid-β deposition (in standardized uptake value ratios; derived from ^18^F florbetapir positron emission tomography) across patients with aMCI and AD from the DMAP cohort. The heatmap in (b) indicates the strength of the alignments between cortical deposition of amyloid-β (x-axis) and the normative neurochemical gradients (y-axis) in these patients. Colors indicate the t-values of these relationships, with asterisks indicating statistical significance after correcting for multiple comparisons. Line plots in (b) show the participant-level simple slopes representing the alignments from (a), with line color indicating whether patients were in the amnestic mild cognitive impairment or probable Alzheimer’s disease clinical subgroup. Cortical maps below are the dot product of the amyloid-β values and neurochemical gradient densities per region, with warm and cool colors indicating positive and negative agreement, respectively, between the two measures. Representative topographies of amyloid-β deposition from patients with high and low alignment to the fourth neurochemical gradient are shown in Figure S4. \*\*\**p*_FDR_ < .005.

### Amyloid-β deposition mediates the alignment of beta rhythmic alterations with cholinergic systems in patients with aMCI and AD

Next, we examined whether any of the alignments of rhythmic neurophysiological alterations with neurochemical systems were mediated by the deposition of Aβ in the DMAP participants. Of the seven alignments, only one exhibited evidence of a potential mediation effect: the relationship between beta rhythmic alterations and the fourth neurochemical gradient (*t* = -2.65, *p* = .008) became non-significant (*t* = -1.84, *p* = .067) when participant-level maps of Aβ deposition were added as a covariate. Causal mediation analysis confirmed that this was a full mediation (Figure 4), as indicated by a significant indirect effect (average causal mediation effect: -0.029, *p* = .018) and a non-significant direct effect (average direct effect: -0.024, *p* = .098). This indirect effect was also significant when specifically considering alignments to muscarinic acetylcholine M1 receptors (average causal mediation effect: 0.01, *p* = .020).

**Figure 4.**
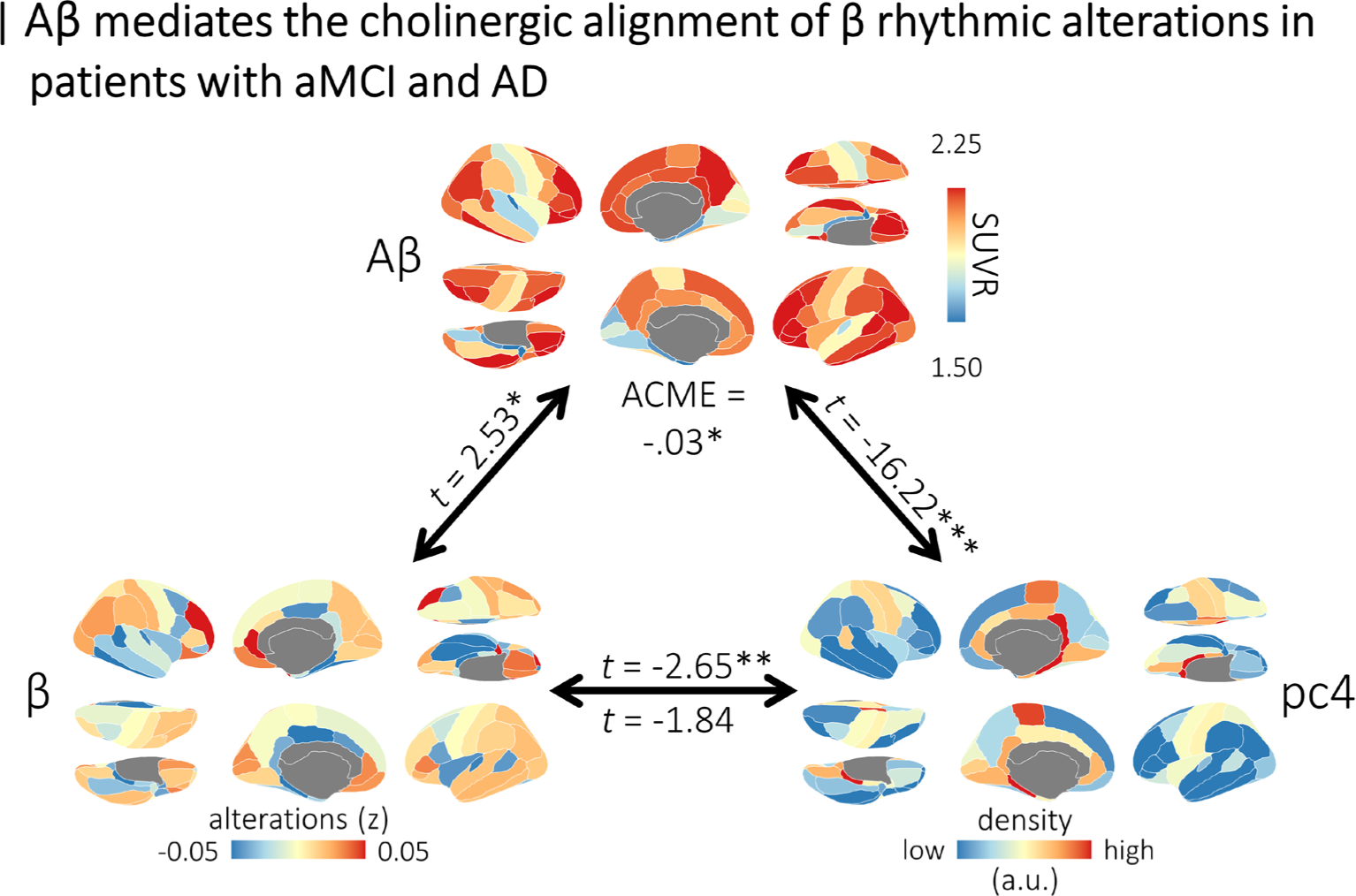
Amyloid-β deposition mediates the neurochemical alignment of beta rhythmic alterations in patients with aMCI and AD. Cortical maps indicate the topographies of mean beta rhythmic alterations (bottom left), mean amyloid-β deposition (top), and the fourth normative neurochemical gradient (bottom right). Linear mixed-effects model statistics (*t*-values) above the paths between these variables indicate their alignment, with asterisks representing statistical significance. The *t*-value below the path on the bottom indicates the residual alignment of beta rhythmic alterations with the fourth neurochemical gradient when amyloid-β deposition is included in the model. The average causal mediation effect (ACME) is shown at the top, confirming an indirect effect. This indirect effect was also significant when the fourth neurochemical gradient was replaced with the atlas of muscarinic acetylcholine M1 receptor densities. \*\*\**p*_FDR_ < .005, \*\**p*_FDR_ < .01, \**p*_FDR_ < .05.

### Neurophysiological and proteinopathic neurochemical alignments are detectable in the asymptomatic stages of Aβ deposition

Finally, we investigated whether neurochemical alignments observed in the later stages of the AD continuum are also detectable in an independent sample of asymptomatic older adults expressing high cortical deposition of Aβ from the PREVENT-AD cohort (Figure 5a). Despite differences in MEG instrumentation, data collection paradigm & site, data processing, and the Aβ PET tracer used (see *Methods: Positron Emission Tomography data collection & processing* and *Magnetoencephalography data collection & processing*), we found that many of the alignments identified in the DMAP participants were detectable in asymptomatic participants with high Aβ from PREVENT-AD (Figures 5b and 5c). Alignments between beta rhythmic alterations and M1 receptors (*t* = 5.42, *p*_FDR_ < .001), and between alpha rhythmic alterations and dopaminergic (DAT: *t* = -9.06, *p*_FDR_ < .001; D1: *t* = -6.91, *p*_FDR_ < .001), glutamatergic (NMDA: *t* = -8.81, *p*_FDR_ < .001), and serotonergic (5-HT1a: *t* = -2.77, *p*_FDR_ = .009; 5-HTT: *t* = -3.50, *p*_FDR_ < .001) systems were replicated in the asymptomatic stages of Aβ deposition.

**Figure 5.**
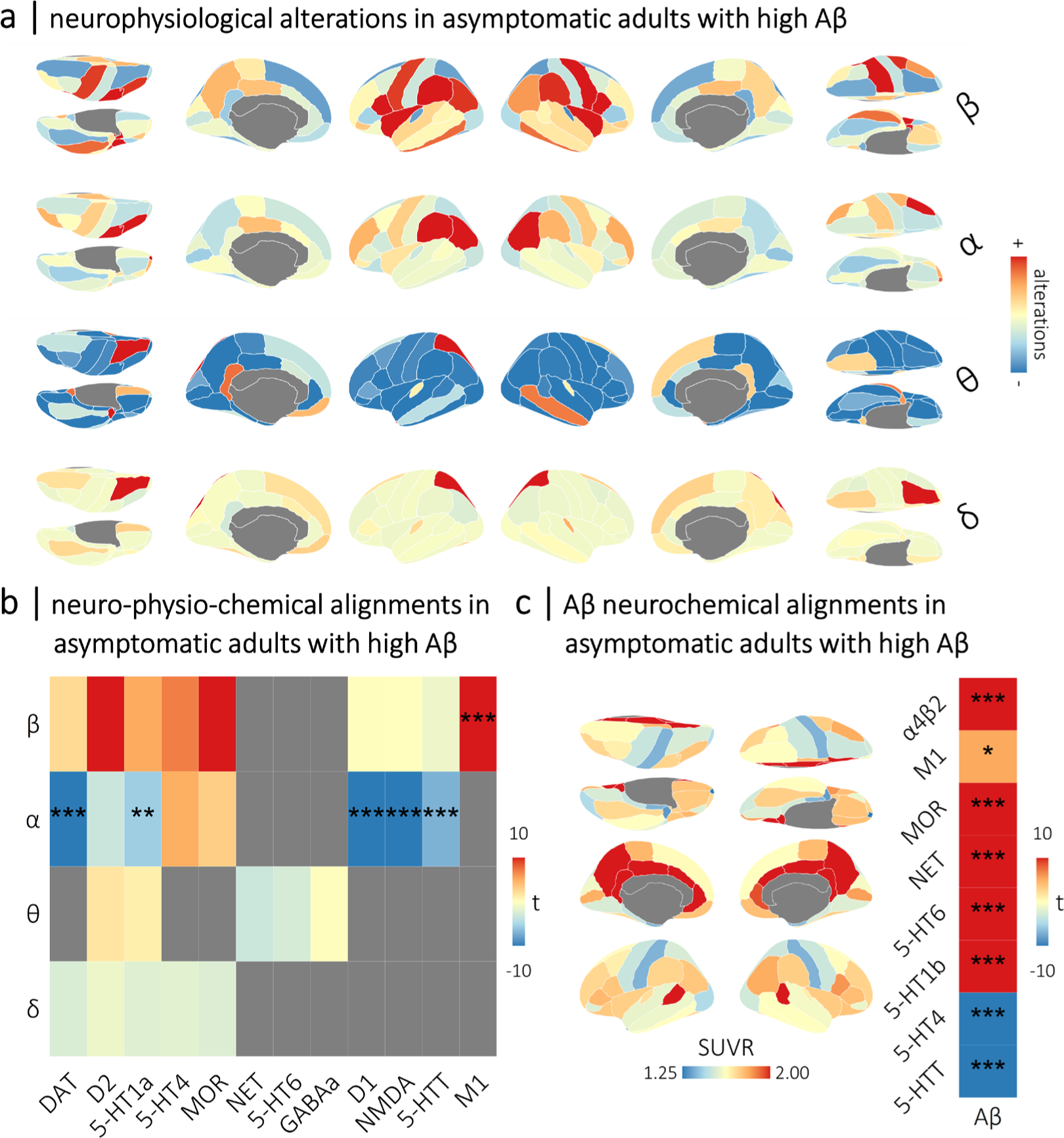
Alignments between normative neurochemical gradients, neurophysiological alterations, and proteinopathy are detectable in the asymptomatic stages of Aβ deposition. The rhythmic neurophysiological measures from the high-Aβ individuals (N = 33) from the PREVENT-AD cohort were standardized to the comparable data from the low-Aβ individuals (N = 71) to generate the neurophysiological alteration maps shown in (a). The heatmap in (b) indicates the replication in PREVENT-AD high Aβ participants of neurochemical-neurophysiological alignments that were observed in patients with aMCI and AD from the DMAP cohort, with band-limited alterations in rhythmic neurophysiology on the y-axis and normative neurochemical atlases on the x-axis. The colors of the squares indicate the t-values of these alignments, with asterisks indicating statistical significance after correcting for multiple comparisons. Grey squares were not significant in the DMAP cohort, and were not examined. Brain maps in (c) indicate the mean topography of Aβ deposition (in standardized uptake value ratios; derived from ^18^F NAV4694 positron emission tomography) across asymptomatic adults with high Aβ deposition from the PREVETN-AD cohort. The heatmap in (c) indicates the replication in PREVENT-AD high Aβ participants of neurochemical-Aβ alignments that were observed in patients with aMCI and AD from the DMAP cohort. Similar to (b), colors indicate the t-values of these relationships, with asterisks indicating statistical significance after correcting for multiple comparisons. \*\*\**p*_FDR_ < .005, \*\**p*_FDR_ < .01, \**p*_FDR_ < .05.

Alignments between regional Aβ deposition and cholinergic (α4β2: *t* = 12.92, *p*_FDR_ < .001; M1: *t* = 2.58, *p*_FDR_ = .012), mu-opioid (MOR: *t* = 6.24, *p*_FDR_ < .001), noradrenergic (NET: *t* = 9.48, *p*_FDR_ < .001), and serotonergic (5-HT6: *t* = 11.90, *p*_FDR_ < .001; 5-HT1b: *t* = 11.03, *p*_FDR_ < .001; 5-HT4: *t* = -7.62, *p*_FDR_ < .001; 5-HTT: *t* = -5.39, *p*_FDR_ < .001) systems were also replicated in asymptomatic individuals with high Aβ (Figure 5c).

We did not find any moderations of the detected neurochemical-neurophysiological alignments by cognitive abilities in these asymptomatic participants (all *p*’s > .05).

## Discussion

Despite evidence of dysfunction across multiple neurotransmitter systems in AD, how this dysfunction shapes cortical neurophysiology has remained unclear. Here, we find that rhythmic, but not arrhythmic, alterations in cortical neurophysiology in patients with aMCI and AD are aligned with the spatial topographies of cholinergic, serotonergic, and dopaminergic systems. The strength of these alignments is associated with the severity of clinical impairments. Marked beta-band increases in muscarinic acetylcholine receptor M1-dense regions are associated with worse attention & executive functions, and stronger delta- and theta-band increases in brain regions rich in dopaminergic, serotonergic, and nicotinic acetylcholinergic systems are related to worse behavioral symptoms. These neurochemical systems also shape the topography of Aβ deposition in patients with aMCI and AD, which accounts for the alignment of beta-rhythmic alterations with cholinergic M1 receptors.

Muscarinic cholinergic M1 receptors emerged in our findings as a particularly important system for shaping the topography of neurophysiological changes and proteinopathy in AD. Increases in beta-rhythmic activity were found to be strongest in M1-dense cortical regions in our study, and this alignment was accounted for by individual variations in the deposition of Aβ plaques. M1 receptors are G-protein coupled cholinergic receptors with the highest density in prefrontal and temporal cortices^71,72^. They promote increased signal-to-noise and synaptic plasticity^72^. M1 receptor dysfunction is tightly linked to AD pathology: Aβ interferes with M1 signaling^23^ and M1 receptor activity enhances non-amyloidogenic processing of amyloid precursor protein^73–75^.

Beta-frequency alterations have also been extensively reported in patients with AD^5,11,76^ and associated with healthy muscarinic cholinergic neuromodulation^77–79^. We found that cholinergic-beta alignments are related to the severity of attention & executive function impairments in patients with aMCI and AD. Reduced function of muscarinic acetylcholine receptors has been tied to attention & executive function impairments in healthy subjects^80–82^ and AD^83^ due to accelerated cytotoxicity. We found that M1 receptor-beta alignments are also expressed in asymptomatic older adults with high levels of cortical Aβ deposition. This finding is promising for novel approaches to pre-clinical pharmacotherapies and disease monitoring. These results unify disparate lines of research, highlighting the association between Aβ deposition, M1 receptor dysfunction, and changes in beta rhythmic neurophysiology in AD, with relevance for cognitive symptoms.

We also found that increased low-frequency (i.e., delta and theta) rhythmic activity in patients with aMCI and AD is expressed along the topographies of dopaminergic and serotonergic cortical systems, and that such alignments are related to the severity of behavioral symptoms. Previous studies reported that dopaminergic and serotonergic functions are related to behavioral and psychiatric issues in patients on the AD continuum^26–29^. However, the neurophysiological bases of behavioral and psychiatric symptoms in AD have, so far, been understudied. Limited previous findings suggest that low-frequency neurophysiological alterations are related to such symptoms^5,84–87^, which our present results confirm. We emphasize, however, that such alignment of low-frequency activity with dopaminergic/serotonergic cortical systems were not present in our data from participants in the asymptomatic stage of Aβ deposition. We may hypothesize that low-frequency neurophysiological activity may not be aligned with these systems in the pre-clinical stage because behavioral symptoms tend to appear later than cognitive impairments in the course of AD^88,89^.

We also report that the deposition of Aβ proteinopathy aligns with the distinct cortical topographies of neurochemical systems in patients with aMCI and AD. Specifically, we found that amyloid plaques are deposited preferentially in regions rich in cholinergic, noradrenergic, serotonergic, mu-opioid, and cannabinoid neurotransmitter systems. Previous work has also linked acetylcholine, serotonin, and norepinephrine dysfunction to Aβ^90^. In our study, we found that only the alignment to cholinergic receptors is related to alterations in neurophysiology. This observation is compatible with the known early susceptibility of cholinergic receptors to Aβ cytotoxicity^91^. The topographical alignment between Aβ deposits and cholinergic, noradrenergic, mu-opioid, and serotonin systems also replicated in our data from asymptomatic participants with early Aβ deposition. Despite these intriguing findings, future investigations using the larger sample of clinical, MRI, and PET data (including both Aβ and tau maps) from the Alzheimer’s Disease Neuroimaging Initiative^92^ are a clear next step for this line of research.

Using normative neurotransmitter atlas data to contextualize cortical alterations in clinical populations has inherent limitations. Foremost of these limitations is the use of PET data collected from healthy participants to contextualize alterations seen in a patient group known to exhibit changes in the same neurotransmitter systems. However, we argue that this aspect only constrains the discussion of our findings, not their robustness. Despite reductions in the local densities of several neurochemical systems in AD, it is unlikely that their respective macro-scale cortical topographies are entirely altered in patients. For this reason, the discussion of our present observations is limited in *how* these systems causally affect neurophysiological alterations and proteinopathy (e.g., whether these changes result from the degeneration of the system or from its compensatory over-activation). Yet, our data can valuably determine *whether* these systems are associated with the pathological processes of the disease. In some cases, this limitation can be overcome by the inclusion of measures known to indicate pathology, such as Aβ deposition. Nevertheless, the *neuromaps* neurotransmitter atlases are currently the only option for performing neurochemical contextualization studies across multiple neurochemical systems. Future research into inter-participant cholinergic, dopaminergic, and serotonergic variability in AD will help fill the remaining knowledge gaps.

Collectively, the present data highlight that neurophysiological alterations and the deposition of harmful proteins in AD co-localize with specific cortical neurochemical systems. These topographical alignments are relevant for hallmark behavioral and cognitive symptoms of the disease, and can be detected even in the asymptomatic stages of Aβ deposition. Looking forward, we anticipate that these findings will inspire further research to validate the mechanistic pathways identified in AD between dopamine and serotonin signaling, low-frequency rhythmic alterations, and behavioral symptoms; as well as between Aβ deposition, muscarinic cholinergic signaling, beta-frequency rhythmic alterations, and cognitive impairments. More detailed and sensitive analyses of the neurochemical alignments of AD Aβ and tau proteinopathies in individual patients are also warranted.

## Materials and Methods

### Participants

Data were aggregated from two studies to examine the clinical (i.e., patients with amnestic mild cognitive impairment [aMCI] and probable AD) and pre-clinical (i.e., asymptomatic older adults with high Aβ deposition) stages of the AD continuum.

### Patients with amnestic mild cognitive impairment and probable Alzheimer’s disease

Data from patients with aMCI and probable AD were included via the Dynamic Mapping of Alzheimer’s disease Pathology (DMAP) study^5^. The Institutional Review Board at the University of Nebraska Medical Center reviewed and approved this investigation, and all research protocols complied with the Declaration of Helsinki. Written informed consent was obtained from each participant (as well as, for participants in the patient group, from their spouse/child informant) following detailed description of the study. For individuals with diminished capacity to make an informed decision regarding research participation, educated assent was acquired from the participant, in addition to informed consent of their legally authorized representative.

Forty-four participants were referred from a memory disorders clinic and screened for recruitment into the aMCI/AD group. All such participants were determined as having either aMCI or mild probable AD by a fellowship-trained neurologist using standard clinical criteria^31^. A positive whole-brain quantitative Aβ PET scan was also required for inclusion into the final aMCI/AD patient participant sample (see *Florbetapir ^18^F Positron Emission Tomography* below). One participant was excluded from this group due to a major incidental MRI finding that was likely to impact cognitive function, and another disenrolled due to COVID-19 related health concerns. Four additional participants were excluded after they were indicated as being Aβ-negative based on their Aβ PET scan. After exclusions, 38 Aβ-positive participants remained for inclusion into the aMCI/AD patient group (aMCI: N = 18, probable AD: N = 20).

For comparison and standardization of the aMCI/AD patient group relative to an analogous group of healthy control participants, 20 additional older adults who reported no subjective cognitive concerns were screened for inclusion into the study. Nineteen of these participants were confirmed to be Aβ-negative by means of a PET scan within the past five years, while one participant received no such test, but performed exceedingly well on all neuropsychological tests. The 19 Aβ-negative participants were recruited based on their previous enrollment in an unrelated clinical trial of an anti-amyloid drug in cognitively healthy older adults, where they were discovered to be Aβ-negative during the screening process and excluded from participation. These participants did not exhibit any cognitive disturbances on detailed neuropsychological assessments.

Exclusionary criteria for both groups included any medical illness affecting CNS function, diagnosis of any neurological disorder (other than Alzheimer’s disease), history of head trauma, moderate or severe depression (Geriatric Depression Scale ≥ 10), and current substance abuse. Demographic factors were matched across the HC and aMCI/AD participant groups (highest level of education: *t* = 1.44, *p* = .156; sex: χ² = 0.84, *p* = .360) with the exception of age (*t* = 2.02, *p* = .048), such that patients with aMCI/AD were younger than those in the HC group. Note that age was included (alongside highest level of education) as a nuisance covariate in all statistical analysis.

### Asymptomatic older adults with high Aβ

Data from asymptomatic older adults with elevated familial risk of sporadic AD were included via the Pre-symptomatic Evaluation of Experimental or Novel Treatments for Alzheimer’s Disease (PREVENT-AD) cohort^32^. The Institutional Review Board at McGill University reviewed and approved this investigation, and all research protocols complied with the Declaration of Helsinki. Written informed consent was obtained from each participant following detailed description of the study. Normal cognition at time of enrollment was assessed using the Montreal Cognitive Assessment (MoCA) and the Clinical Dementia Rating scale (CDR). Individuals with MoCA scores < 27 or CDR scores > 0 were further examined by a neuropsychologist from the PREVENT-AD group to verify their cognitive status. Individuals with neurological or psychiatric illness affecting CNS function were excluded. From the subsample of 124 participants who underwent both Aβ PET and resting-state MEG, 20 were excluded due to issues with data quality (N_MEG_ = 19, N_PET_ = 1). The remaining 104 participants were then divided into high-and low-Aβ individuals based on an established sensitive threshold (see *Materials and Methods: Positron Emission Tomography*; high-Aβ: N = 33; low-Aβ: N = 71). The asymptomatic low-and high-Aβ groups did not significantly differ in age (*t* = -0.36, *p* = .721), sex (χ² = 1.37, *p* = .241), or highest level of education (*t* = 0.94, *p* = .352). These demographics were also matched between the high-Aβ group from PREVENT-AD and the patients with aMCI/AD from DMAP on age (*t* = -1.06, *p* = .293) and education (*t* = -1.34, *p* = .185), but not sex (χ² = 7.40, *p* = .007).

Demographics for each group from the DMAP and PREVENT-AD studies, as well as comparisons between groups, can be found in Table 1.

**Table 1.**
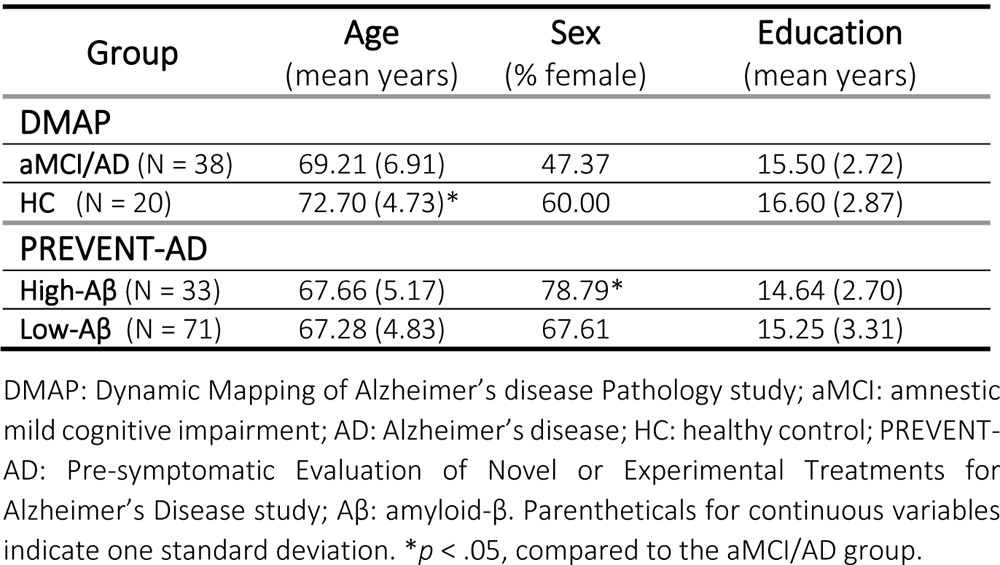
Group demographics and comparisons.

### Neuropsychological & clinical testing

Participants in the DMAP study completed a series of neuropsychological tests focusing on five cognitive domains: attention & executive function, memory, learning, verbal function, and processing speed. Raw scores for each participant were converted to demographically adjusted z-scores based on published normative data^33–36^ and averaged within each functional domain to create composite domain z-scores. Details on the neuropsychological tests administered, as well as on the statistical independence of the domain z-scores can be found in previous publications^5,37,38^. In collaboration with a spouse or child informant for patients with aMCI and AD, instrumental activities of daily living (iADLs) were measured using the Functional Activities Questionnaire (FAQ)^39^, and dementia severity and behavioral symptoms were measured using the Quick Dementia Rating System (QDRS)^40^. General cognitive status was measured using the MoCA^41^ and the Mini-mental State Examination (MMSE) ^42^.

Participants in the PREVENT-AD study underwent cognitive assessment using the Repeatable Battery for the Assessment of Neuropsychological Status (RBANS)^43^, which yields scaled scores subdivided into five cognitive domains comprising immediate and delayed memory, attention, visuospatial constructional abilities, and language.

We focused our analyses on the cognitive domains most commonly associated with AD: attention & executive function and memory^44^. We also examined associations with IADLs and behavioral symptoms (measured via the behavioral symptom subscale of the QDRS, in the DMAP study only).

### Positron emission tomography data collection & processing

In the DMAP study, combined PET/CT data using ^18^F-florbetapir (Amyvid™, Eli Lilly) were collected following procedures described by the Society of Nuclear Medicine and Molecular Imaging (3D acquisition; single intravenous slow-bolus < 10 mL; dose = 370 MBq; waiting period = 30-50 min; acquisition = 10 min)^45^ using a GE Discovery MI digital PET/CT scanner (Waukesha, WI). PET images were attenuation-corrected using the CT data, and reconstructed in MIMNeuro (slice thickness = 2 mm)^46^, converted to voxel-wise standardized uptake values based on body weight (SUVbw), and then normalized into MNI space. Each scan was read by a fellowship-trained neuroradiologist, who was blinded to their group assignment, and assessed as being “Aβ-positive” or “Aβ-negative” using established clinical criteria^46^. Those who were Aβ-negative were excluded from the patient group at this point. Images were then normalized to the crus of the cerebellum (SUIT template)^47^ to generate voxel-wise maps of SUV ratios^48^, and back-transformed into native space using each individual’s FreeSurfer-processed T1 MRI data. The PET data overlapping with each individual’s cortical gray-matter ribbon was projected onto a tessellated *FSAverage* template surface using mri_vol2surf (maximum value; projection fraction = 1; steps of 2)^49^, and spatially smoothed (FWHM: 8mm)^50,51^. These data were then averaged within each region of the Desikan-Killiany atlas^52^.

In the PREVENT-AD study, PET imaging data were collected using a Siemens HRRT head-only, high-resolution PET camera^32^. Aβ scans were performed 40 to 70 minutes after injection of ≈6mCi of ^18^F-NAV4694 (Navidea Biopharmaceuticals, Dublin, OH). Images were reconstructed using a 3D OP OSEM (10 iterations, 16 subsets) algorithm, and were decay and motion corrected. Scatter correction was performed using a 3D scatter estimation method^53^. PET images were preprocessed using an in-house pipeline from the Villeneuve lab (https://github.com/villeneuvelab/vlpp). Briefly, the 4D image files (6 frames of 5 minutes for NAV) were realigned, averaged, and registered to their corresponding structural MRI. Standardized uptake value ratio (SUVR) maps were generated by using the cerebellum gray matter as a reference region for Aβ scans. The resulting SUVR values were averaged across each parcel from the Desikan-Killiany atlas^52^ and then again over a meta-ROI of early Aβ-accumulating cortical regions^54^ and converted them to centiloid scale following established procedures and formulas^55,56^. Participants were divided into high-and low-Aβ groups based on a centiloid threshold of 18 (SUVR = 1.249), which is optimal for predicting future cognitive decline in asymptomatic individuals^57,58^.

### Magnetoencephalography data collection & processing

In the DMAP study, participants were seated in a nonmagnetic chair with their head positioned within the MEG sensor array, and rested with their eyes closed for 8 minutes^59^. Recordings were conducted in a one-layer magnetically-shielded room with active shielding engaged for environmental noise compensation. Neuromagnetic responses were sampled continuously at 1 kHz with an acquisition bandwidth of 0.1– 330 Hz using a 306-sensor Elekta/MEGIN MEG system (Helsinki, Finland) equipped with 204 planar gradiometers and 102 magnetometers. Preceding MEG measurement, four head position indicator coils were attached to the participant’s head and localized, together with the three fiducial points and about 100 scalp surface points, using a Fastrak 3-D digitizer (Polhemus Navigator Sciences, Colchester, VT, USA). Each MEG dataset was individually corrected for head motion and subjected to noise reduction using the signal space separation method with a temporal extension (correlation limit: .950; correlation window duration: 6 seconds) ^60^. Only data from the gradiometers were used for analysis. Structural MRI data were also collected for each participant, for co-registration of their MEG data (Siemens Prisma 3T; 64-channel head coil; TR: 2.3 s; TE: 2.98 ms; flip angle: 9°; FOV: 256 mm; slice thickness: 1 mm; voxel size: 1 mm^3^).

In the PREVENT-AD study, at least two 5-minute runs (i.e., a total of 10 minutes)^59^ of resting state, eyes open MEG data were collected from the participants^59^ as they fixated on a central crosshair displayed on a screen. Data were collected using a 275-channel whole-head CTF system (Port Coquitlam, British Columbia, Canada) at a sampling rate of 2400 Hz and with an antialiasing filter with a 600 Hz cut-off. Built-in third-order gradient compensation was applied to attenuate environmental noise. About 100 scalp points were localized in each participant with a Polhemus Fastrak device, including anatomical landmarks at the nasion and the left and right preauricular points. Head movements during MEG data collection were monitored with head position indicators attached to the participants’ forehead and the left and right mastoids, alongside reference signals for heartbeats and eye movements with concurrent electrocardiographic and electrooculographic recordings. Structural MRI data were also collected for each participant, for co-registration of their MEG data (Siemens TIM Trio 3T; 12- or 32-channel head coil; TR: 2.3 s; TE: 2.98 ms; flip angle: 9°; FOV: 256x240x176 mm; slice thickness: 1 mm; voxel size: 1 mm^3^).

For both studies, triangulated cortical surfaces were derived from the T1 MRI data with *FreeSurfer* recon_all^49^ using default settings and imported into *Brainstorm* ^61^. Using digitized points of the scalp and fiducials, each participant’s MEG data were co-registered with their own high-resolution structural T1-weighted MRI data using an iterative closest-point rigid-body registration in *Brainstorm* and manually corrected following visual inspection. Individual cortical surfaces were down-sampled to 15,000 vertices for use in MEG source imaging.

*Brainstorm* was used for MEG data preprocessing and analysis and followed recommended good-practice procedures^62^. After import into *Brainstorm*, MEG data were filtered (DMAP: bandpass = 1 - 200 Hz & notch = [60, 120, 180] Hz; PREVENT-AD: high-pass = 0.3 Hz & notch = [60, 120, 180, 240, 300] Hz), and ocular and cardiac artifacts were identified using an automated identification algorithm, supplemented by visual inspection of their temporal and spatial topography. Signal-Space Projectors (SSPs) were generated for each type of artifact, the temporal and spatial topography of these SSPs were reviewed, and those accounting for ocular and cardiac components were removed from the gradiometer data. Artifact-reduced MEG data were then arbitrarily epoched into non-overlapping blocks of 4 s and, in the DMAP study, downsampled to 500 Hz. Epochs still containing major artifacts (e.g., SQUID jumps) were excluded within each participant using the ∪ of standardized thresholds median absolute deviations from the median (DMAP: ± 2.5 MAD; PREVENT-AD: ± 3.0 MAD) for peak-to-peak signal amplitude and gradient. Empty-room recordings of ≥ 2 min, collected around each individual scanning session, were processed using an identical pipeline to the one described above (with the exception of the artifact SSP), to model the environmental noise statistics for source analysis.

In both studies, MEG data were source imaged using an overlapping-spheres forward model. For the DMAP study, a linearly constrained minimum variance beamformer implemented in *Brainstorm* was used to spatially-filter the epoch-wise data with source orientations unconstrained to the cortical surface. In the PREVENT-AD study, we estimated the imaging kernels using depth-weighted dynamic statistical parametric mapping (dSPM) with cortical current flows oriented perpendicularly to the cortex^63^. The source-level time series data were then transformed into the frequency-domain using Welch’s method (DMAP: window = 1s; 50% overlap; PREVENT-AD: window = 2s; 50% overlap) and parameterized using *specparam*^64^ (Brainstorm Matlab version; DMAP frequency range = 2 – 30 Hz; PREVENT-AD frequency range = 1 – 40 Hz; Gaussian peak model; peak width limits = 0.5 – 12 Hz; maximum n peaks = 3; minimum peak height = 3 dB; proximity threshold = 2 standard deviations of the largest peak; fixed aperiodic; no guess weight).

To represent the arrhythmic features of the neurophysiological power spectrum, the slope and offset of the aperiodic model fit were extracted at each vertex. The rhythmic (i.e., aperiodic-corrected) spectra were derived by subtracting the aperiodic spectra from the original PSDs. Arrhythmic and rhythmic neurophysiological features were averaged over vertices within each region of the Desikan-Killiany atlas^52^. Rhythmic spectral data were then averaged over canonical frequency bands (delta: 2–4 Hz; theta: 5–7 Hz; alpha: 8–12 Hz; beta: 15–29 Hz). This procedure produced six maps of neurophysiological brain activity per participant: one for rhythmic activity in each of the four canonical frequency bands, and one for each of the broadband arrhythmic model features (i.e., aperiodic slope and offset).

### Normative atlases of neurotransmitter system density

Similar to previous work^30^, we used *neuromaps*^65^ to obtain mean cortical distribution maps of 19 receptors and transporters, following previously-established procedures^66^ and parcellated the resulting topographies according to the Desikan-Killiany atlas^52^. We obtained normative densities for dopamine (D1: 13 adults, [11C]SCH23390 PET; D2: 92, [11C]FLB-457, DAT: 174, [123I]-FP-CIT), serotonin (5-HT1a: 36, [11C]WAY-100635; 5-HT1b: 88, [11C]P943; 5-HT2a: 29, [11C]Cimbi-36; 5-HT4: 59, [11C]SB207145; 5-HT6: 30, [11C]GSK215083; 5-HTT: 100, [11C]DASB), acetylcholine (α4β2: 30, [18F]flubatine; M1: 24, [11C]LSN3172176; VAChT: 30, [18F]FEOBV), GABA (GABAa: 16, [11C]flumazenil), glutamate (NMDA: 29, [18F]GE-179; mGluR5: 123, [11C]ABP688), norepinephrine (NET: 77, [11C]MRB), histamine (H3: 8, [11C]GSK189254), cannabinoids (CB1: 77, [11C]OMAR), and opioids (MOR: 204, [11C]Carfentanil). We derived the principal patterns of spatial variance (i.e., gradients) across these 19 maps using principal component analysis (PCA, with each map first scaled and centered), using the *prcomp* and *PCAtest*^67^ functions in *R*^68^. Permutation testing was used to determine statistically-significant principal components^67^, with *p*-values calculated by comparing the empirical eigenvalue of each component to a null distribution of eigenvalues derived from 1,000 random permutations of the underlying data. We retained the principal components with *p* < .05 for further analysis, as representative topographies of the principal normative cortical neurochemical gradients of the human brain. To test the importance of synapse density, we also retrieved the cortical atlas topography of synaptic vesicle glycoprotein 2A (76 adults, [11C]UCB-J) from *neuromaps*.

### Modeling of spatial colocalization

To quantify the topographies of neurophysiological alterations for each participant on the AD continuum (i.e., patients with aMCI/AD from DMAP, asymptomatic adults with high Aβ from PREVENT-AD), the neurophysiological features were z-scored with respect to the means and standard deviations of analogous data from participants without high Aβ deposition (i.e., the healthy control group from DMAP, the low-Aβ participants from PREVENT-AD).

We estimated the alignment of neurophysiological alterations and Aβ deposition with the *neuromaps* principal gradients using linear mixed-effects modeling^30^ via the *nlme* package in *R*. For each participant on the AD continuum, this approach models a nested intercept and slope representing the linear relationship between these region-wise cortical features and the selected *neuromaps* gradient, and thus exploits the within-subject variability in the data that would be ignored by group-level colocalization analysis. These models were computed with the following form: *neuromap pc ∼ alterations + covariates, random = (∼1 +* alterations | *participant)*. Significant relationships between neurochemical gradients and alterations were displayed by plotting the nested simple slopes between these variables per participant.

We examined whether significant neurochemical alignments are moderated by cognitive function (i.e., attention & executive function, memory), IADLs (i.e., FAQ scores; available only in the DMAP study), and behavioral symptoms (i.e., QDRS-Behavioral scores; available only in the DMAP study) by including these variables as interaction terms in the linear mixed-effects models. The effects of significant continuous moderators were visualized by extracting the nested model slope coefficients per participant and plotting them against the moderator. We also tested significant alignments in the DMAP participants for moderations by clinical subgroup (i.e., aMCI versus probable AD).

For each significant neurochemical alignment, as well as for significant moderations of these alignments, we performed post-hoc testing to assess the specificity of the statistical effect to each neurotransmitter system. This was done by regressing (i.e., using a comparable linear mixed-effects model) the relevant alterations on each *neuromap* atlas that exhibited significant loadings onto the relevant neurochemical gradient.

All models included age and highest level of education as cross-sectional nuisance covariates. We used the Benjamini-Hochberg method to correct parametric tests for multiple comparisons across related hypotheses, with a threshold for significance set to *p_FDR_*< .05.

Initial step-wise linear mixed-effects models were used to identify potential mediation effects^69^. Indirect effects were then tested statistically via causal mediation analysis with quasi-Bayesian approximation using the *mediation* package in *R*^70^.

## Data availability

Due to concerns over patient confidentiality the MEG and PET data from the DMAP study cannot be shared on an open repository, but will be made available from the corresponding author upon reasonable request.

Some of the data from the PREVENT-AD study are publicly available (https://openpreventad.loris.ca/, https://registeredpreventad.loris.ca/ and https://www.mcgill.ca/bic/neuroinformatics/omega)^32^. The remaining data can be shared upon approval by the scientific committee at the Centre for Studies on Prevention of Alzheimer’s Disease (StoP-AD) at the Douglas Mental Health University Institute.

Normative neurotransmitter density data are available from *neuromaps* (https://github.com/netneurolab/neuromaps).

## Acknowledgements

We would like to acknowledge the efforts of our research participants, and thank them for their selflessness and kind demeanour. We would also like to thank the research and clinical staff who conducted patient recruitment and data collection for this study.

This work was supported to AIW by a Banting Postdoctoral Fellowship from the Canadian Institutes of Health Research (CIHR; BPF-186555) and grant F32-NS119375 from the United States National Institutes of Health (NIH); to JGR from the Mexican National Council of Science and Technology (CONACyT; 2020-000017-02EXTF-00402) and the Healthy Brains Healthy Lives program at McGill University; to SV from the Alzheimer Society of Canada, the Alzheimer Association, the Canadian Institutes of Health Research (CIHR, 438655), and a CIHR Canada Research Chair Tier 2 and a Brain Canada Platform grant; to SB from a NSERC Discovery grant (RGPIN-2020-06889), the CIHR Canada Research Chair (Tier 1; CRC-2017-00311) of Neural Dynamics of Brain Systems, a grant from the NIH (R01-EB026299), and by the Canada Brain Research Fund (CBRF), an innovative arrangement between the Government of Canada (through Health Canada) and Brain Canada Foundation, and Alzheimer’s Association; and to TWW from the NIH (R01-MH116782-S1; R01-MH118013-S1). The funders had no role in study design, data collection and analysis, decision to publish, or preparation of the manuscript.

PREVENT-AD was launched in 2011 as a $13.5 million, 7-year public-private partnership using funds provided by McGill University, the FRQS, an unrestricted research grant from Pfizer Canada, the Levesque Foundation, the Douglas Hospital Research Centre and Foundation, the Government of Canada, and the Canada Fund for Innovation. Private sector contributions are facilitated by the Development Office of the McGill University Faculty of Medicine and by the Douglas Hospital Research Centre Foundation (http://www.douglas.qc.ca/).

## Author Contributions

AIW, SV, SB and TWW contributed to the conception and design of the study; AIW and JGR contributed to the analysis of the data; and AIW contributed to drafting the text and preparing the figures. All authors contributed to editing of the manuscript.

## Potential Conflicts of Interest

The authors report no conflicts of interest.

## Notes

### Competing Interest Statement

The authors have declared no competing interest.

### Author Declarations

Data from patients with aMCI and probable AD were included via the Dynamic Mapping of Alzheimer's disease Pathology (DMAP) study. The Institutional Review Board at the University of Nebraska Medical Center reviewed and approved this investigation, and all research protocols complied with the Declaration of Helsinki. Written informed consent was obtained from each participant (as well as, for participants in the patient group, from their spouse/child informant) following detailed description of the study. For individuals with diminished capacity to make an informed decision regarding research participation, educated assent was acquired from the participant, in addition to informed consent of their legally authorized representative. Data from asymptomatic older adults with elevated familial risk of sporadic AD were included via the Pre-symptomatic Evaluation of Experimental or Novel Treatments for Alzheimer's Disease (PREVENT-AD) cohort. The Institutional Review Board at McGill University reviewed and approved this investigation, and all research protocols complied with the Declaration of Helsinki. Written informed consent was obtained from each participant following detailed description of the study.

