## Supplementary Materials for "Alignments between cortical neurochemical systems, proteinopathy and neurophysiological alterations along the Alzheimer’s disease continuum"

Alex I. Wiesman* et al.

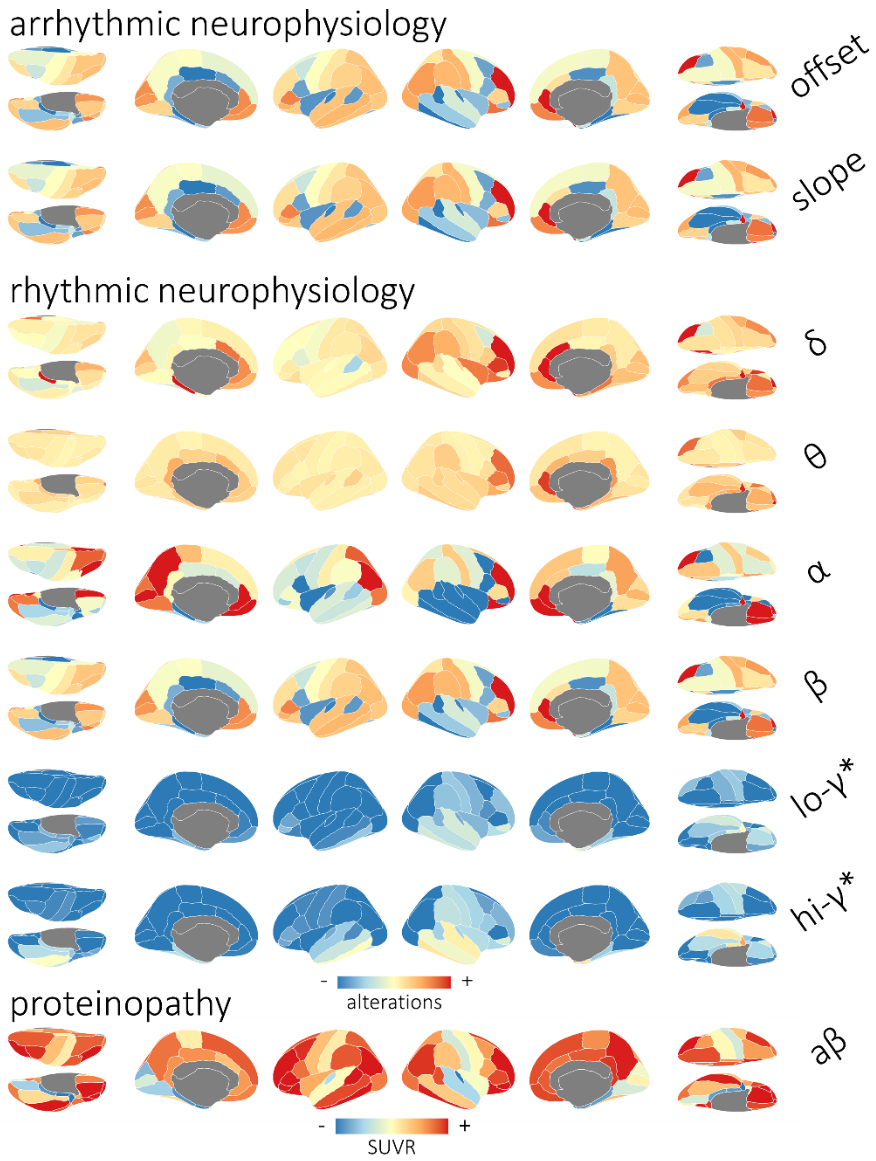


**Figure S1. Topographies of neurophysiological alterations and proteinopathy in patients with aMCI and AD.** Cortical maps indicate the group-average spatial topographies of alterations in arrhythmic (top) and rhythmic (middle) neurophysiology in patients with aMCI and AD from the DMAP cohort. Note that these topographies represent neurophysiological measures that have been standardized (i.e., z-scored) to the comparable data from the healthy control participants. Warm and cool colors indicate increases and decreases in rhythmic activity relative to healthy levels, respectively. Cortical maps at bottom indicate the mean spatial topography of amyloid-β deposition (in standardized uptake value ratios; derived from $^{18}$F florbetapir positron emission tomography) across these same patients.

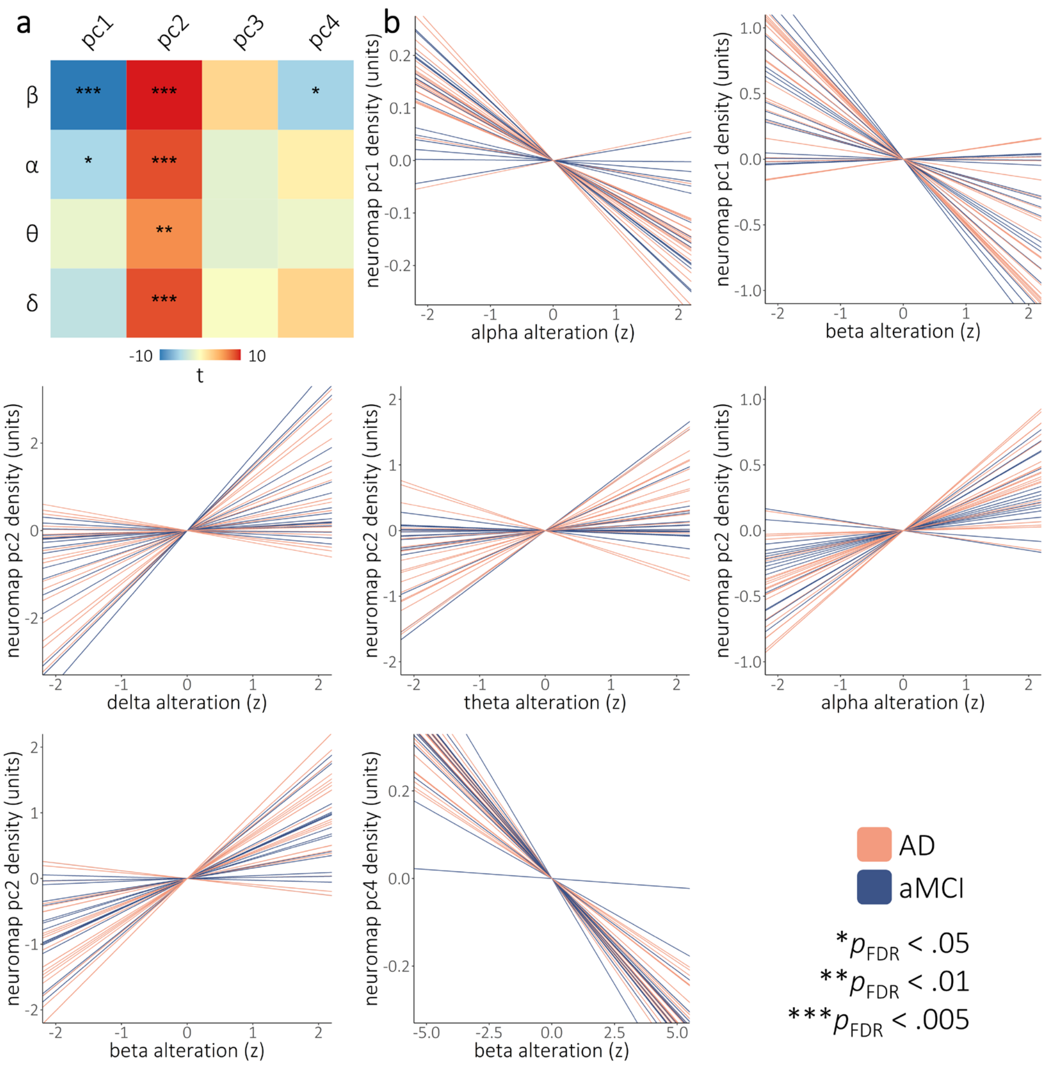


**Figure S2. Neurophysiological alterations in patients with aMCI and AD align to neurochemical systems.** Similar to Figure 2, the heatmap in (a) indicates the strength of the alignments between band-limited alterations in rhythmic neurophysiology (y-axis) and the normative neurochemical gradients (x-axis) from Figure 1. Colors indicate the t-values of these relationships, with asterisks indicating statistical significance after correcting for multiple comparisons. Line plots in (b) show the participant-level simple slopes representing all such significant alignments from (a). Line colors indicate whether patients were in the amnestic mild cognitive impairment or probable Alzheimer's disease clinical subgroup. ***$p_{FDR} < .005$, **$p_{FDR} < .01$, *$p_{FDR} < .05$.

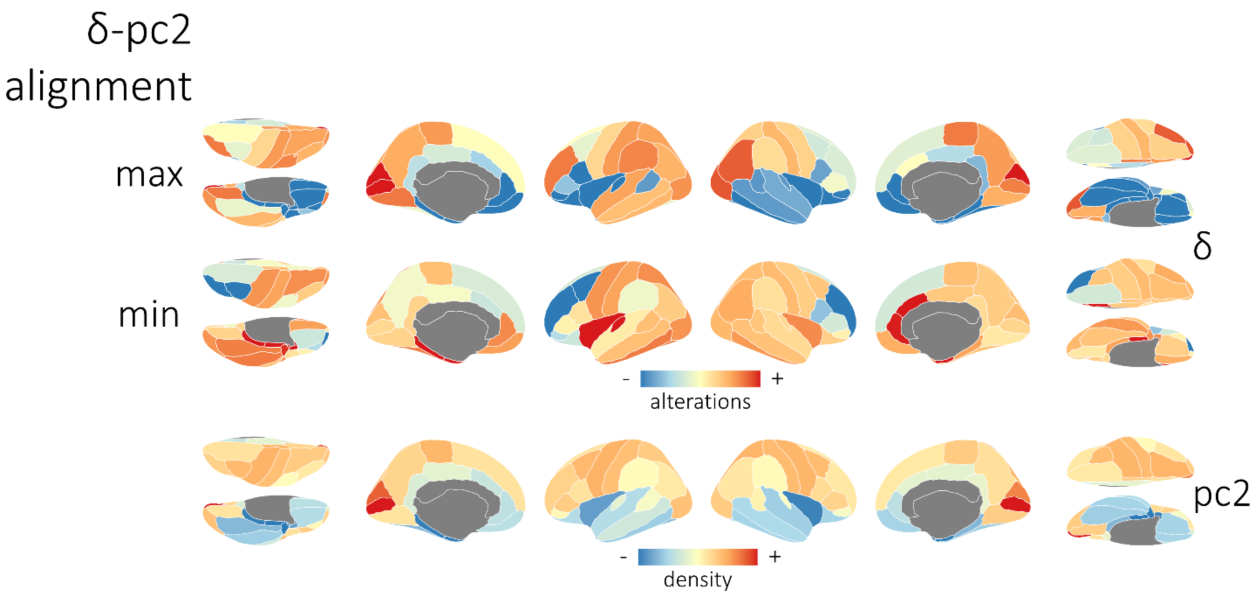


**Figure S3. Representative topographies of delta rhythmic alterations from aMCI/AD patients with high and low alignment to the second neurochemical gradient.** Cortical maps above indicate the topographies of delta rhythmic alterations for the representative DMAP patients with the strongest (i.e., most positive) and weakest (i.e., least positive) alignment of these data to the topography of the second neurochemical gradient (below).

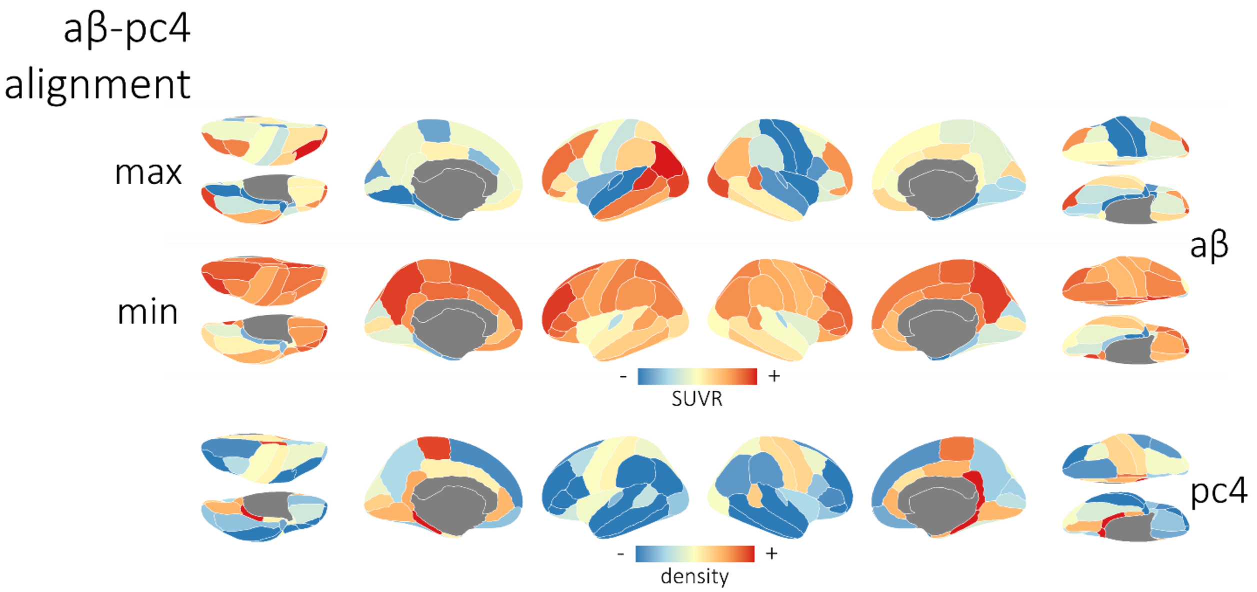


**Figure S4. Representative topographies of amyloid-β deposition from aMCI/AD patients with high and low alignment to the fourth neurochemical gradient.** Cortical maps above indicate the topographies of amyloid-β deposition for the representative DMAP patients with the strongest (i.e., most negative) and weakest (i.e., least negative) alignment of these data to the topography of the fourth neurochemical gradient (below).
